# Building sustainable local training capacity in maternal and newborn health within the public system-A training intervention research in Palghar District, western Maharashtra, India

**DOI:** 10.1101/2024.01.01.24300686

**Authors:** Shilpa Karvande, Vidula Purohit, Subha Sri Balakrishnan, Helen Allott, Elisabeth Serle, Reeta Jha, Shubhro Mullick, Milind Chavan, Prashant Kulkarni, Matthews Mathai, Nerges Mistry

## Abstract

**Introduction:** Constant investment for professional development of health workforce is paramount and can be implemented through locally available competent trainers. However, guidelines for selection and recruitment of trainers, and for approaches and processes of training and nurturing them are lacking. A training intervention research in western Maharashtra, India focusing on maternal and newborn health (MNH) attempted to address these gaps.

**Methods:** During the pilot project in Pune District (2016-19), 38 health workers were trained as trainers in MNH with support from the United Kingdom (UK) based technical partners (obstetricians and pediatrician). From these, eight were selected and further trained to become core trainers (CTs) for MNH training during scale-up phase in Palghar District (2019-22). A local cadre of skilled master trainers (MTs) in Palghar (n=32) was developed by these CTs with support from the UK team, through a six-steps approach starting with pre-training of trainers (TOT) screening, TOT with pre– and post-training assessment, staggered induction of MTs, continuous support, regular quality assessment, and virtual refresher training. A mixed-methods approach was adopted to assess the process and outcome of this six-step approach.

**Results:** The six-steps approach of developing Palghar MTs helped in optimal resource utilization during TOT. It allowed more time and non-threatening support to MTs to excel through peer learning. Pune CTs became competent and confident mentors through critical feedback and appreciation and provided long term support to the newly trained Palghar MTs. Regular assessment by the UK team was an essential quality control step. The virtual learning served as an effective mode for refresher training without interruptions during COVID-19. Considerable improvement in clinical knowledge and skills of the trainee health providers from Palghar (n=505) was evidence of quality training delivered by the Palghar MTs. The training intervention contributed to increased clinical competency, confidence, recognition, motivation, and contribution to career growth of the Palghar MTs.

**Discussion:** The TOT model used in Palghar demonstrated the importance of recruiting right candidates with essential attributes as trainers, conscious shift in their teaching style for making them adult-learner-centric facilitators, longer time frames and hybrid platform. A methodical approach for nurturing talent of trainers through opportunities for teaching as well as learning was highlighted. However, a systematic evaluation for assessing long term effects of this model on retention of clinical and teaching competencies of these trainers and their trainees and its application in provision of MNH care is required.

## INTRODUCTION

A competent public health workforce is an important pillar of health system for improving population health.^1,2^ However, constant intensive efforts are required to maintain its competencies. Pre-service training lays the foundation of competencies but regular refreshing and updating are essential to maintain them. Therefore, the need to invest in in-service training and continuous professional development of the health workforce is paramount. To be effective, training programs must focus on appropriate training techniques, provided by competent faculty possessing clinical and teaching skills.^3–5^ Moreover, the need for urgent transformation in training approaches is evident, as traditional didactic techniques with passive instructions e.g. lectures have little or no impact on learning outcomes.^6^ Case-based learning, clinical simulations, skills practice, and feedback are more effective training techniques of participant engagement and show positive learning outcomes in terms of increased knowledge, skills, and confidence while delivering healthcare.^6–8^

A training cascade is a process intended to strengthen workforce capacity at various levels. A ‘Training of Trainer’ (TOT) model works on the premise of the right selection of professional trainers who have the ability to acquire, understand, and transfer knowledge and skills to trainees or peers and to sustain the same for continuous use in future.^9,10^ Various studies have shown the effects of the TOT model in implementing evidence-based public health practices.^11–15^ This model is considered as a strong predictor of sustainability because of its potential for up-skilling the workforce rapidly, cost-effectively, continuously, and exponentially.^10^ Furthermore, qualified trainers can act as health facility-based mentors to continually reinforce the lessons learnt during routine practice.

In addition to essential clinical knowledge and skills, non-technical attributes for trainers of TOT include good communication skills, time management, team work, and respectful behaviour.^16^

However, evidence-based guidelines for selection and recruitment of trainers for public health training programs are limited in India and research studies are needed in this area. Similarly, evidence for pedagogies and processes of public health training using a TOT model is lacking.

In this context, a training intervention research was undertaken in western Maharashtra focusing on maternal and newborn health (MNH). It involved a TOT model and was implemented as a pilot project in Pune District (2017-19) and later scaled up to Palghar District (2019-22). The present paper describes the pedagogy, process, and effect of the TOT model in Palghar district on developing a resource of sustainable and competent trainers for MNH in-service training within the local public health system.

## METHODS

### Study setting

The study was conducted in Palghar District in western Maharashtra. This district has a population of 2,990,116^17^ with majority of tribal communities in six of its eight rural blocks. Low birthweight babies, teenage pregnancies, and anemia are some of the high-risk factors related to MNH in Palghar.^18,19^ Due to these reasons, the Government of Maharashtra recommended this district for the scale-up phase of the project.

The primary health care system of the district consists of 46 Primary Health Centers (PHCs) and 311 Health Sub Centers (HSCs). A PHC, the first port of call to a qualified doctor in the public sector, typically covering a population of 30,000 in plain areas and 20,000 in hilly, tribal, or difficult areas. It acts as a referral unit for HSCs.^20^ An HSC is the most peripheral and first point of contact between the primary health care system and the community covering a population of 5000 in the plains and 3000 in tribal and hilly areas for provision of essential preventive and promotive and few curative and referral services.^20^

### Study design

An intervention research for building capacity of local trainers and health workforce for MNH was implemented in Pune (13 rural blocks) during 2016-2019 ^8^ as a pilot project and scaled up in Palghar district during 2019-2022 (at a time when India experienced three waves of the COVID-19 pandemic). Both these projects, were implemented with on-site (for Pune) and virtual (for Palghar) support from the technical partners-a team of obstetricians and pediatrician-from the United Kingdom (UK). The intervention research project design involved two phases-a) Preparing for the scale-up in Palghar and b) Developing a local cadre of skilled master trainers in Palghar

### Phase 1-Preparing for the scale-up in Palghar

A group of eight clinically competent trainers-seven General Nurse Midwives (GNMs) and one medical doctor-(out of 38) from the Pune pilot project were selected as the ‘Core’ trainers (CTs). These were the ones who could imbibe and repeatedly demonstrate expected training pedagogy; and expressed willingness and commitment to work in the scale-up phase in Palghar. They had received continuous and critical feedback on clinical and teaching skills from the UK team during the pilot phase. Acknowledging the seriousness of the prevalent health issues in Palghar district, the scale-up in Palghar included two major adaptations in the training content. Firstly there was an addition of or greater emphasis to local context specific subjects related to MNH e.g. management of low birth weight babies, anemia, and malnutrition. Secondly it targeted additional cadres of health providers. The Pune pilot project had focused only on auxiliary nurse midwives (ANM) and there was a felt need of training an entire team of health providers (medical officer, staff nurse, community health officer, GNM and ANM) working at PHC/ HSC since MNH case management would generally be a team work. The Pune pilot project’s training material, as well as the adapted content for Palghar was reviewed and endorsed by experts from the State Family Welfare Bureau, Government of Maharashtra.

The Pune CTs were oriented about the scale-up phase in Palghar and their potential roles. A four-day refresher learning session followed by a series of virtual continuous learning sessions, each focusing on one key subject related to MNH, were organized for them over a period of three months. These sessions included their lectures using PowerPoint presentations and clinical skill demonstrations with critical feedback provided by the UK team experts to sharpen their clinical knowledge, practical skills and teaching skills as CTs. A specific session was conducted by the UK experts about “*How to become an effective trainer*?” with practical tips and guidance for the Pune CTs.

### Phase 2-Developing a local cadre of skilled master trainers in Palghar

The approach of developing a local cadre of skilled master trainers (MTs) for Palghar district involved six steps-

#### Step 1-Pre-TOT screening of identified candidates by Pune CTs and project researchers

A group of 48 candidates (medical officers, staff nurses, community health officers, obstetricians, pediatricians working as clinicians and/or administrators at various levels of health facilities in the District) were identified as potential trainers. This identification was based on the field-based interactions with Palghar district health officials, health providers and observations by the project researchers during initial exploratory visits and baseline assessment. A pre-TOT screening of these identified potential TOT candidates over four days focused primarily on the assessment of their work profile, previous experience as trainer, if any, interest and commitment to work as a trainer, and demonstration of teaching skills. The teaching skill demonstration was assessed for a) session preparations, b) clarity in conveying messages, c) ability to engage participants, and d) being respectful during teaching. They were scored using a three-point grade scale-excellent, fair, and not satisfactory. The candidates who received ‘excellent’ or ‘fair’ grades were considered as potential TOT participants (n=32; 25 medical doctors and seven GNMs.

#### Step 2-Training of potential master trainers with pre– and post-TOT assessment

The 32 selected candidates participated in the TOT to become Palghar MTs, with pre– and post-TOT assessments of clinical knowledge and skills, and teaching skills. Adult-learner-centric principles such as respecting participants’ previous experience, ensuring their active engagement and hands-on practice with explanation of step-wise management, etc. were applied during the TOT and all the subsequent trainings. A simulation of various cases with high risks/ complications related to MNH was the approach used for discussing the step-wise case management. The pre– and post-TOT assessment included written examination, structured observation of clinical demonstration using mannequins and teaching skills with a check-list. Of the 32 Palghar MTs, 18 participants achieved a composite score of more than 85% and the remaining 14 scored in the range of 70-84% in the post-TOT assessment.

#### Step 3-Staggered induction of selected candidates as MTs

A strategic process of staggered and structured induction was adopted for 32 Palghar MTs for conducting training of health providers from the public health system in Palghar. MTs with ‘intermediate’ scores (70-84%) (n=14) were paired up with those with the ‘highest’ scores (85% and above) (n=18). Those with intermediate scores were inducted as independent trainers to train health providers after satisfactorily working with the ‘highest’ scoring ones in at least two training batches. The Palghar MTs conducted training of 505 health providers over a period of 18 months (Table 1).

**Table 1.** – Cadre-wise profile of health providers (n=505) trained by Palghar MTs.

|  |  |
| --- | --- |
| **Cadre** | **No. of trained health providers** |
| Auxiliary Nurse Midwife (ANM) | **304** |
| Community Health Officer (CHO) | **73** |
| Medical Officer | **66** |
| General Nurse Midwife (GNM) | **38** |
| Lady Health Visitor (LHV*) | **18** |
| Staff nurse | **6** |
| **TOTAL** | **505** |

\* Lady Health Visitor (LHV) is an Auxiliary Nurse Midwife who is trained for a period of six months to function as a female health supervisor and provides supportive supervision and technical guidance to ANMs
(https://main.mohfw.gov.in/sites/default/files/Training%20Division.pdf)

#### Step 4-Continuous support and guidance to Palghar MTs by Pune CTs

Each training batch of Palghar health providers (n=20-25) was conducted by a mix of ‘highest’ and ‘intermediate’ scoring Palghar MTs (altogether 4-5) along with the support from one Pune CT. Beyond clinical guidance and critical feedback, this supportive supervision included teaching them training pedagogy for confirming adherence to adult-learner-centric principles such as being participatory, interactive and respectful, exhibiting positive learning approach, giving opportunities for participants to think and self-learn, and ensuring adequate time for reverse demonstration by each participant. (Reverse demonstration-The trainer demonstrated a clinical skill to a group of 5-6 participants using a mannequin in a stepwise manner and asked each participant to observe. Subsequently each of them was expected to demonstrate it to the trainer, engaging rest of the participants, and the trainer observed and provided feedback to them.) This step provided longer time frame of working as a trainer under supportive supervision for the Palghar MTs.

#### Step 5-Regular quality assessment by the UK experts

Due to travel constraints during the Covid-19 pandemic, structured quality assessment and critical feedback sessions (n=3) were conducted by the UK experts virtually. These sessions helped in ensuring correct demonstration of clinical skills and standardized delivery of training content by the Palghar MTs. While these Palghar MTs had the essential clinical knowledge, they found it challenging to teach case-based sessions and giving opportunity to trainees to think and learn. They were provided with feedback and tips in the initial rounds of teaching demonstrations for adapting to this pedagogy. Most of them immediately took corrective steps as suggested by the UK experts and showed improvements in the subsequent rounds of skill demonstrations. Examples include-encouraging participants to think about next steps in case management, exercising respectful communication throughout the case-based discussions and correct demonstration of abdominal palpation during pregnancy.

#### Step 6-Virtual refresher training for MTs

Though an intense one-time face-to-face training can lead to increased knowledge and clinical skills, there is a risk of attrition over a period of time. Moreover, face-to-face training is both resource and time intensive. Repeating the same over time was not feasible especially during the COVID-19 pandemic. Acknowledging this reality, a series of refresher sessions on essential MNH skills were organized by the project team for the Palghar MTs with support from the Pune CTs. These sessions were conducted over a period of 10 months (starting six months after the TOT), using ECHO-Extended Community Health Outcomes India (a virtual knowledge-sharing platform for building capacity and sharing best practices through case-based learning using a hub and spoke model.^21^ These refresher sessions were planned on the principle of learning through discussions of real-life clinical situations presented by the Palghar MTs. The essential skills included plotting and interpretation of partograph for monitoring progress of labor, management of eclampsia, newborn resuscitation and abdominal palpation of pregnant women.

### Data collection and analysis

To assess the process and outcomes of this methodical six-step approach of generating a cadre of local MTs in Palghar, a mixed-methods approach was adopted. The pre– and post-training assessment data were quantitative and included scores assigned for clinical knowledge and skills. These data were entered and analyzed using Microsoft Excel. Descriptive statistics were used to compute and compare the pre– and post-training assessment scores.

The qualitative data were collected through process documentation, structured observations and in-depth interviews of UK clinical experts (n=3), Pune CTs (n=5), and Palghar MTs (n=12) These data were coded and analyzed thematically for key themes such as experience of training pedagogy, effects of training on Pune CTs and Palghar MTs in terms of building confidence and competencies, and application of training in practice.

### Ethics statement

This intervention research project received approval from the Institutional Research and Ethics Committee.

## RESULTS

Results of this training intervention research are presented as a) the importance of the methodical six-step approach of developing Palghar MTs, b) evidence of increased competency of Palghar MTs with support from Pune MTs, c) personal and professional gains of the Palghar MTs

### Importance of the methodical six-step approach of developing Palghar MTs

A group of eight Pune CTs contributed to building a cadre of 32 Palghar MTs-(25 medical doctors and seven GNMs) during the scale-up phase. Each of the six-steps adopted in the approach of building the cadre of Palghar MTs was found to have its own advantages (Table 2).

**Table 2.**
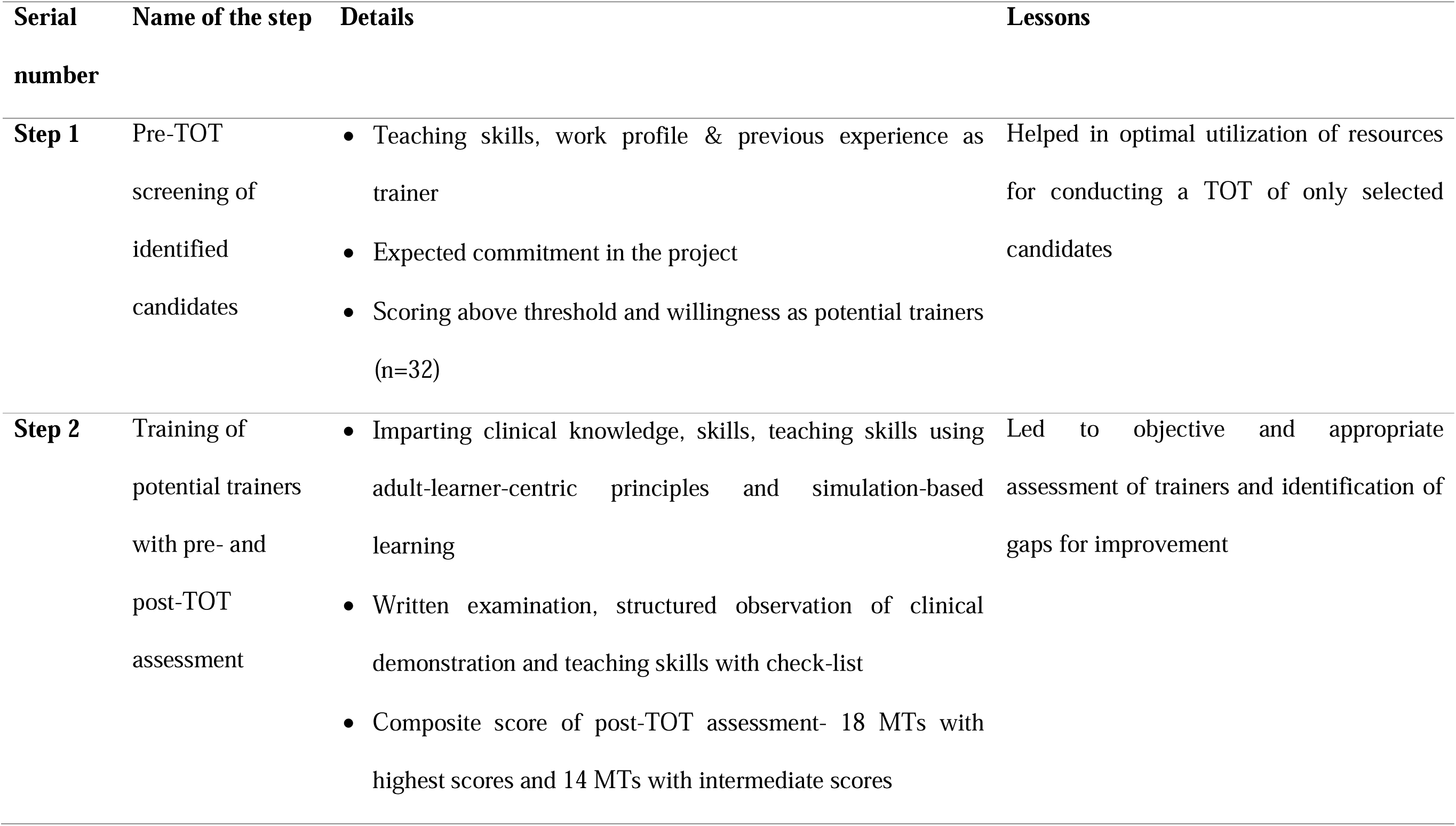
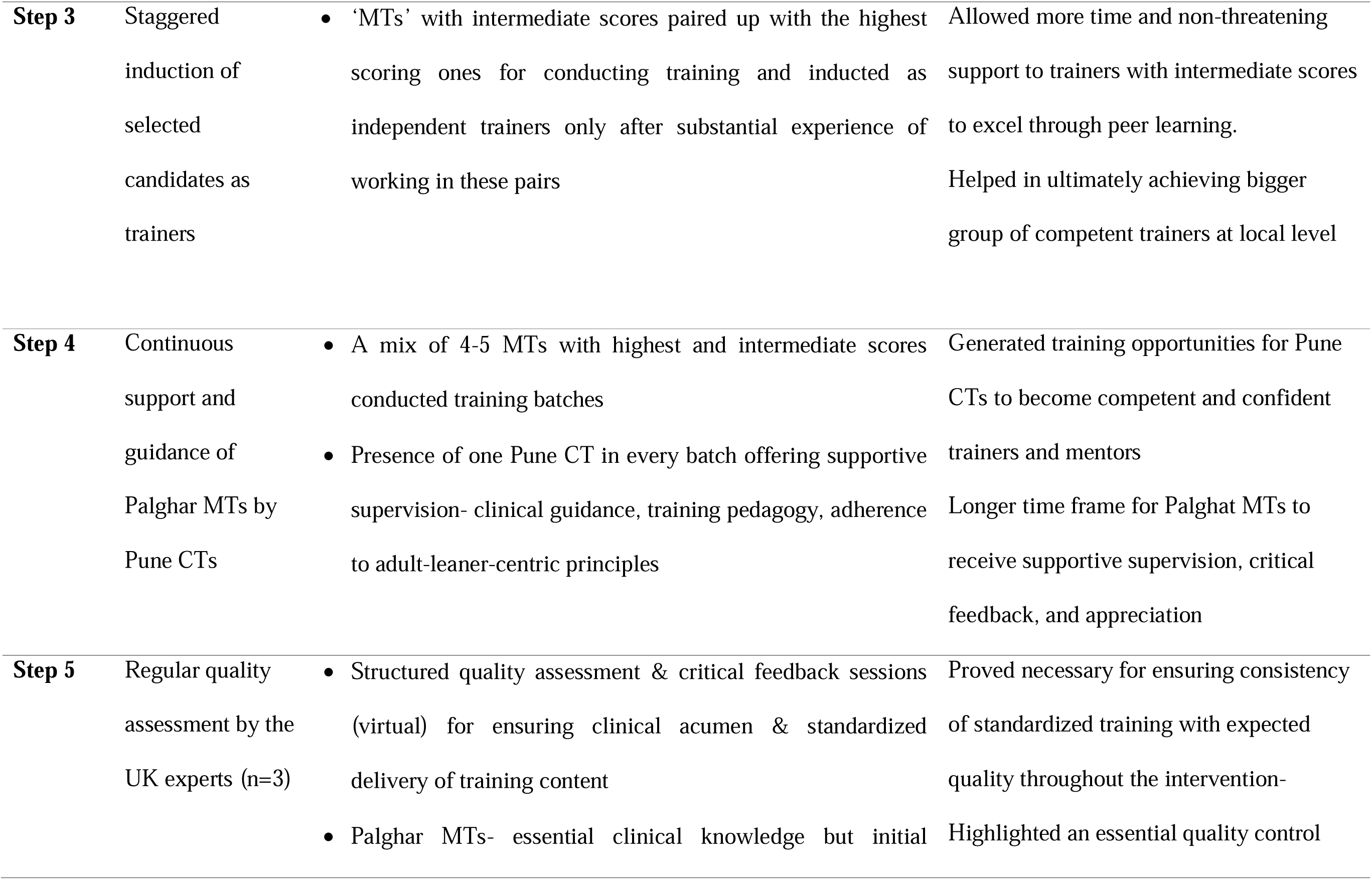
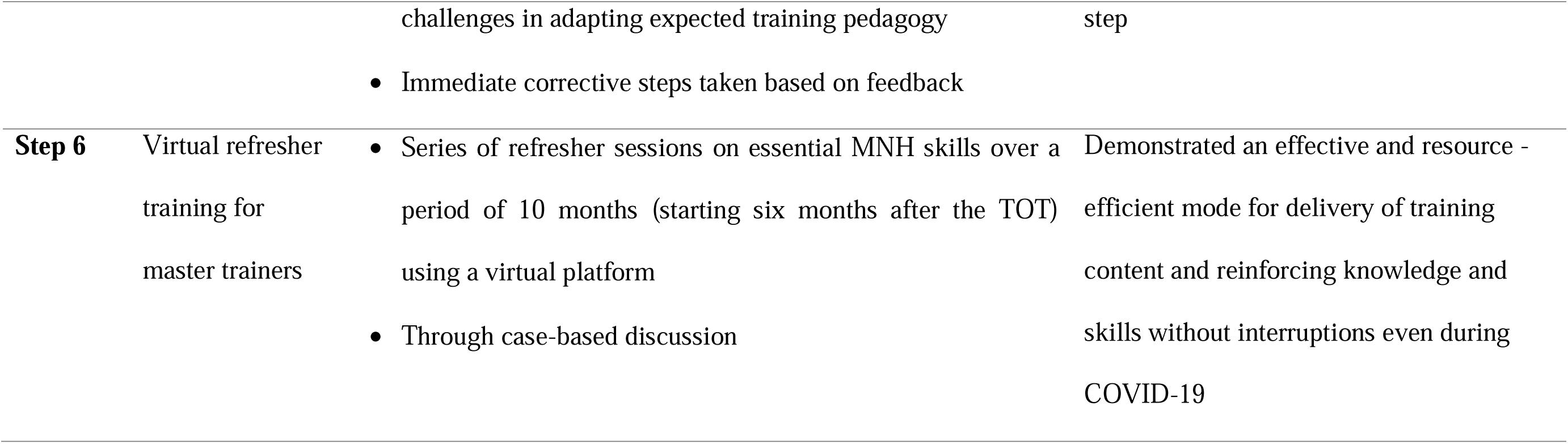
– Methodical six-step approach of building MT cadre and lessons learnt.

### Evidence of increased competency of Palghar MTs with support from Pune MTs

In-person training received from the Palghar MTs and Pune CTs with periodic virtual inputs from the UK experts resulted in considerable improvement in clinical knowledge and skills (abdominal palpation, newborn resuscitation and eclampsia management) of the trainee health providers from Palghar (Fig 1)

**Figure 1.**
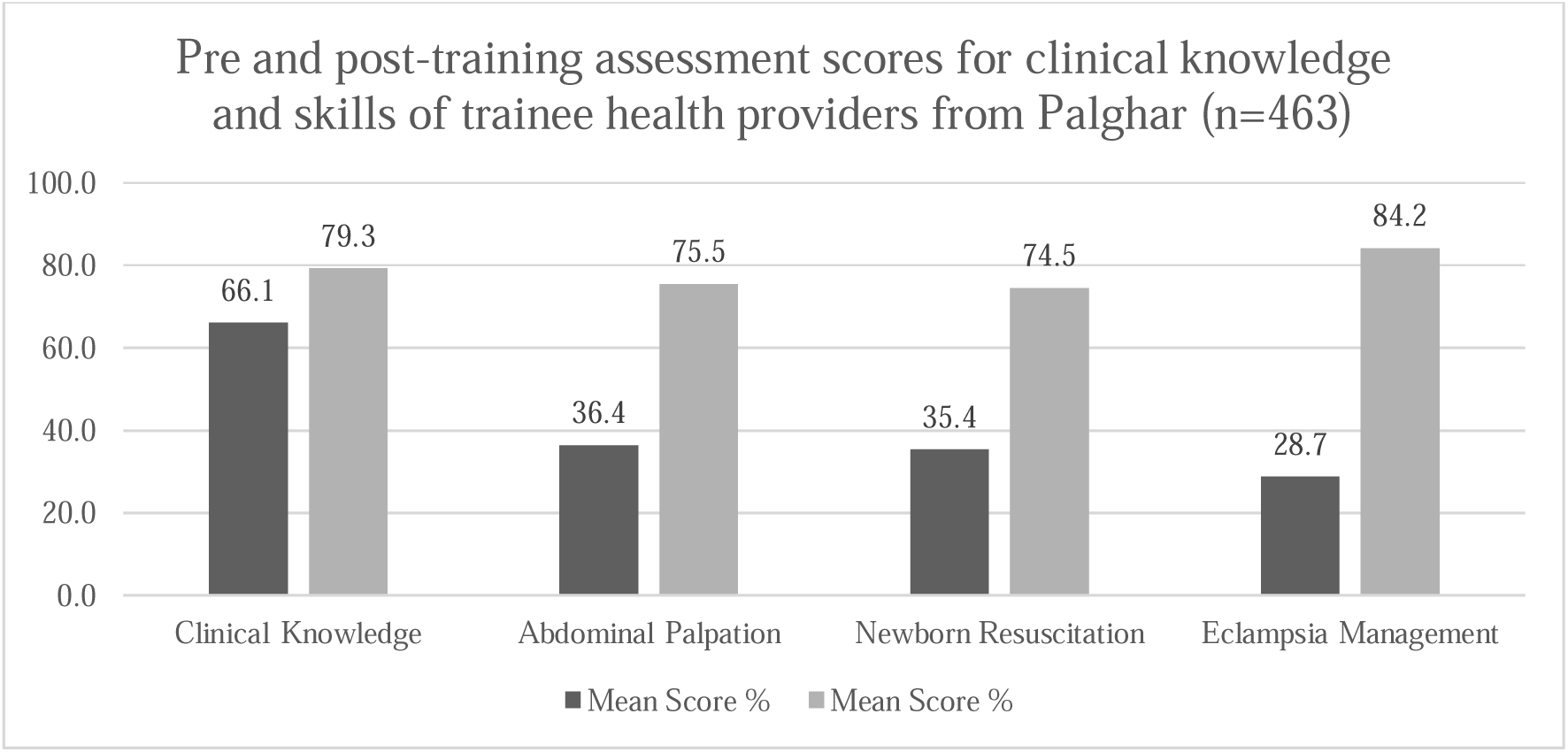
– Pre and post-training assessment scores for clinical knowledge and skills of trainee health providers from Palghar (n=463)

> *(Though 505 health providers were trained during the training intervention, pre– and post-training scores for all the assessments were available only for 463 providers)*

The post training assessment scores were significantly higher than that of the pre-training assessment scores for clinical knowledge (z scores= –17.423, P value-0.000), as well as for the three skills namely-abdominal palpation (z scores=-18.384, P value-0.000), newborn resuscitation (z scores=-18.423, P value-0.000) and eclampsia management (z scores=-18.644, P value-0.000). This improvement in the post-training assessment scores of the trainee health providers was an evidence of the quality of training delivered to them by the Palghar MTs with support from the Pune CTs.

### Personal and professional gains of the Palghar MTs-

The effect of the current training intervention on the Palghar MTs had three aspects-i) increased clinical competency, ii) confidence as a trainer, and iii) career growth-recognition and motivation.

### Increased clinical competency and its application

The Palghar MTs were practicing clinicians-staff nurses, non-specialist or specialist medical doctors. They reported internalizing the clinical protocols thoroughly on account of repeated demonstration of essential MNH skills during the training. They could acquire leadership roles at their health facilities for monitoring and mentoring their staff for appropriate case management and referrals.

> *“When we demonstrated the skills in several training batches, it got fixed in our heads. Now we have become more effective in managing any crucial case.” Palghar MT (Senior specialist, Sub District Hospital (SDH))*
>
> *“I check all the data for my PHC especially all the high-risks and the referred-out data to see whether the required primary management was undertaken here before referring the cases.”-Palghar MT (MO-PHC)*
>
> *“I taught about partograph plotting to all our staff nurses. It helps everyone including myself to understand obstructed labor.” – Palghar MT (MO-Rural Hospital (RH))*

The Palghar MTs appreciated learning new concepts e.g. Danger Response-Airway Breathing Circulation (DR ABC) approach of case assessment which helped them to be more organized and efficient in systematic patient management. The training had emphasized upon appropriate referral practices and use of standardized referral tool-Situation Background Assessment Recommendation (SBAR).^22^ The Palghar MTs’ preparedness to manage referred-in cases increased because of the appropriate referral notes being sent with the patients and because of the appropriate primary-level management undertaken before referral. This reduced delays in secondary management of patients at referral facility and increased chances of their recovery and survival. They also mentioned adapting to evidence-based practices and avoiding harmful practices which were previously followed without assessing their clinical relevance or checking evidence.

> *“Due to this training not only me but also my ANMs started following DR ABC approach very diligently and not missing out important investigation such as measuring respiratory rate.” Palghar MT (CHO)*
>
> *“We were copying our seniors and giving fundal pressure when the baby would be stuck; or rupture the membrane artificially. I learnt about consequences of such practices and have stopped these things. I have even told the same to my colleagues.”-Palghar MT (Staff nurse, RH)*
>
> *“Recently I have received one referral from PHC. The child had pneumonia and was in respiratory distress. The case was not manageable by the PHC staff. They referred the child to our facility but with oxygen. So we could start higher treatment immediately.”-Palghar MT (MO, SDH)*
>
> *“Even when I want to refer a case to a higher hospital, I don’t hesitate to call the specialist and describe the case as per the Situation Background Assessment Recommendation (SBAR) taught to us during this training.” – Palghar MT (MO, RH)*

### Confidence as a trainer

Confidence building of trainers was another focus area in this intervention. The Palghar MTs were engaged as a group in conducting repeated training of health providers. This resulted in more opportunities for peer interactions, encouragement from Pune CTs and the UK experts and increased confidence. With experience of working as MTs in the project, they started applying the newly learnt adult-learner-centric training pedagogy in their own health facilities, in other settings and training programs. Following quotes explained the nuances of increased confidence mentioned by the Palghar MTs –

> *“I wanted to improve my stage daring [way of describing stage fright] hence I participated as a trainer in number of batches during this project. I learned many new things such as how to present a topic or how to organize a training programs. I now feel very confident.” Palghar MT (MO-PHC)*
>
> *“I am sharing my knowledge of case management with my colleagues who could not attend this training. That’s a wonderful feeling which also increases our own knowledge.”-Palghar MT (Staff Nurse, SDH)*
>
> *“Earlier my ‘teaching pattern’ was just to talk and talk; and give information to students who would come and learn nutrition here. I now make their groups, ask them to interact with mothers of admitted children here, conduct a diet audit and make them think about possible reasons for malnutrition among children. I understood how I can involve the students and engage them for learning.”-Palghar MT (MO, Nutrition Rehabilitation Center, SDH; who manages medical college students interning at their hospital)*

### Career growth-recognition and motivation

Apart from the direct benefits to the Palghar MTs in terms of increased clinical competency and confidence as a trainer, the training intervention additionally benefitted them for receiving recognition or appreciation from colleagues and superiors and added value to their leadership roles in their health facilities. They expressed willingness to continue working as a trainer in future and even getting promoted as a CT. Receiving critical feedback as well as appreciation from the UK experts during the live virtual quality assessment sessions was a motivating moment for the Palghar MTs.

> *“My image as a medical officer has changed. Even other PHC doctors call me or discuss with me about any clinical problem because they all know that I work as this project’s trainer. Problem gets solved by sharing, so it benefits.” Palghar MT (MO-PHC)*
>
> *“The enthusiasm of the trainers [Palghar MTs] is worth noting especially moving forward as a group in these challenging years [of COVID pandemic]-UK expert*
>
> *“After receiving this training, even my senior Sir asks for my clinical opinion. I feel good that my knowledge is being useful.” – Palghar MT (MO-RH)*
>
> *“I feel so motivated. Even I would like to work like Pune CT and travel to other place to train more trainers.”-Palghar MT (MO-PHC)*

## DISCUSSION

The MNH and midwifery related skill deficits and hence the urgent need to strengthen the health workforce is evident in various studies.^8,23–25^ The low pre-training assessment scores of health providers from Palghar confirmed the same. Recent initiatives in India such as skills lab, Dakshata training^26^ and India’s National Training Strategy^27^ have shifted focus to skills-based training strategies; however the benefits of these strategies are yet to percolate to frontline health workers from the primary health system. India will face a shortfall of approximately 0.7 million skilled health workers to reach the 25: 10000 ratio of skilled health workers to population by 2030.^25^ In this context, the current training intervention with a training cascade of Pune CTs, Palghar MTs and Palghar health providers demonstrated a model to upskill the existing human resource within the public health system.

This training intervention adopted a TOT model and presented its effects on developing sustainable competent trainers within the public health system. There are several studies that document the content and criteria for assessment of TOT ^28,29^ however little evidence is available about the approach, implementation steps or prerequisites of TOT for achieving expected gains.^10^ The TOT planning and evaluation require capturing outcome as well as process measures, the ultimate aim and the steps required to achieve it.^30^ This paper has attempted to describe the necessary elements of the methodical approach and outcomes of the TOT intervention.

The right choice of trainers is a prerequisite for a successful TOT intervention. Enthusiasm, willingness to teach, patience, insight, confidence, communication skills, leadership, capacity for self-reflection, ability to be constructively critical, and above all motivation to help others are considered to be important attributes of a trainer.^10,31^ The Pune pilot study demonstrated importance of systemic investments in creating and sustaining competent, motivated, and updated trainers and conducive environments for absorption of learning among ANMs.^8^ The Palghar scale-up phase deliberately invested efforts to identify and recruit right candidates with essential attributes for minimizing attrition of skilled resource.^10^

Appropriate training pedagogy is the next essential for effective training intervention. Adult learning process can be transformative and transformational.^28,32–34^ Hence a conscious shift in the teaching style of the trainers was attempted by making them adult-learner-centric facilitators and effectively using simulation-based learning approach which has been valued in training intervention research.^35–37^ The positive outcomes of the current training intervention in terms of increased confidence among Palghar MTs and enhancement in clinical knowledge and skills of health providers demonstrated the appropriateness of this approach. This training intervention built on a hybrid platform with an appropriate blend of virtual and in-person training activities proved to be effective for delivery of training content and reinforcing knowledge and skills even during COVID-19 pandemic. Other studies have validated the usefulness of virtual platform for conducting training during COVID-19 pandemic.^35^

Training of trainers (TOTs) is generally a short-term program with little opportunities for MTs to conduct supervised training.^38^ Longer time frames are needed for trainers to learn, assimilate and then teach others^10^ and newly trained trainers need to be supported beyond the training event ^9^ which gets missed out in a typical one-time TOT event. Gradual and staggered involvement of Palghar MTs and continued support by Pune CTs with periodic guidance from the UK experts provided an extended opportunity to the newly trained Palghar MTs to unlearn and learn, assimilate, accept, and act on constructive criticism, and then conduct training of health providers under supportive supervision.

The Palghar MTs reported increased capacity and motivation in performing their routine tasks. This concept of perceived increased ability to perform tasks successfully is termed as ‘self-efficacy’ and self-efficacy following continuous education is known to increase knowledge and motivation to implement the new knowledge for patient care.^39^ However, self-reporting of gains or positive effects of intervention could be overestimated.^37^ In the present research, the qualitative data from in-depth interviews of and observations from the Pune CTs, UK experts and project researchers endorsed this increased ability Palghar MTs thus minimizing the possibility of overestimation.

Studies have recommended that becoming a trainer should be recognized as a career path and not just an adjunct duty so as to avoid attrition of trainers^10^ and such training programs should be integrated within systemic mechanisms or processes.^40^ Reluctance in releasing staff for training and perceiving it as an unnecessary burden primarily due to limited human resource has been reported in other studies.^10,41^ There was no such reluctance in releasing Palghar MTs for participating in the training. However, at times it was challenging for them to commit the necessary time against their competing clinical and/or administrative priorities. Hence a realistic and wider strategy needs to be developed for creating space for clinicians to work as trainers while making use of their skills continuously. An efficient training intervention with TOT model should have self-sustaining exponential growth and continue generating an ever-widening pool of skilled human resource. However, this swift progression is not possible in the absence of enabling service conditions which may lead to attrition of skilled trainers.^10^ Hence it demands continued investment in human resources for health and enablers in health facilities including embedded refresher training, uninterrupted supply of drugs and materials, and essential infrastructure. There needs to be a systematic approach for nurturing talent of trainers through accreditation and provision of opportunities for teaching as well as learning. The study recommends use of longer time frames, hybrid platform and investment in enabling ecosystem for applying learning, as part of this approach.

This training intervention research has highlighted the benefits of a methodical approach for nurturing talent of trainers through opportunities for teaching as well as learning. However, a systematic evaluation for assessing long term effects of this training intervention on retention of clinical and teaching competency of these trainers and its application in provision of MNH care is further required. Future research for studying costing of such a model of training intervention will be warranted if it is to be scaled up and institutionalized.

## Data Availability

All data produced in the present study are available upon reasonable request to the authors.

